# Using Lambda phages as a proxy for pathogen transmission in hospitals

**DOI:** 10.1101/2022.09.09.22279770

**Authors:** Kylie B. Burke, Brandon A. Berryhill, Rodrigo Garcia, David A. Goldberg, Joshua A. Manuel, Paige Gannon, Bruce R. Levin, Colleen S. Kraft, Joel Mumma

## Abstract

One major concern in hospitalized patients is infections with pathogens borne on surfaces, patients, and healthcare workers. Fundamental to controlling nosocomial infections is identifying the sources of pathogens, monitoring the processes responsible for their transmission, and evaluating the efficacy of the procedures employed for restricting their transmission. Here we present a method using the bacteriophage Lambda (λ) to achieve these ends. Defined densities of multiple genetically marked λ phages were inoculated at known hotspots for contamination on high-fidelity mannequins. Healthcare workers (HCWs) then entered a pre-sanitized simulated hospital room and performed a series of patient care tasks on the mannequins. Sampling occurred on the scrubs and hands of the HCWs, as well as previously defined high-touch surfaces in hospital rooms. Following sampling, the rooms were decontaminated using procedures designed and demonstrated to be effective. Following the conclusion of the simulation, the samples were tested for the presence, identity, and densities of these Lambda phages.

The data generated enabled the determination of the sources and magnitude of contamination caused by the breakdown of established infection prevention practices by HCW. This technique enabled the standardized tracking of multiple contaminants during a single episode of patient care. While our application of these methods focused on nosocomial infections and the role of HCW behaviors in their spread, these methods could be employed for identifying the sources and sites of microbial contamination in other settings.

## Introduction

One of the greatest harms of being hospitalized is infections acquired from pathogenic microbes in treatment environments (e.g., catheter-associated urinary tract infections or catheter-associated bloodstream infections) (1). These pathogens can be unknowingly transmitted by healthcare workers (HCWs) to patients, thereby contributing to patient morbidity and mortality (2).Previous studies using mathematical models (3-5) have shown a solution to reducing the incidence of hospital-acquired infections is to increase the efficacy of measures for preventing the transmission of pathogens from HCWs to patients (e.g., hand hygiene and barrier precautions). Central to designing, implementing, and evaluating these measures is elucidating the sources of the pathogens responsible for infections and the pathways for transmitting those pathogens to patients.

As a patient care environment can harbor different sources of pathogens (6) and abounds in opportunities for their transmission, there is a need to simultaneously trace multiple transmission pathways. Existing approaches using non-biological or biological surrogates for pathogens do not satisfy the need for *in situ* evaluation of transmission events. Non-biological surrogates, such as fluorochrome-tagged body fluids, can be used to simulate different sources of contamination simultaneously but not the susceptibility of pathogens to disinfectants (7). Approaches using biological surrogates, including live viruses and viral DNA markers, are limited. Viruses, such as bacteriophage MS2, are susceptible to disinfectants (8) but can only be used to trace a single source of contamination at a time. Viral DNA markers, such as cauliflower mosaic virus DNA or silica nanoparticles with encapsulated DNA (9), can produce multiple unique markers (8(10) but are unaffected by commonly used disinfectants (such as 70% ethanol; (8)). Thus, a method is needed for tracing multiple transmission pathways simultaneously during patient care, each of which can be counteracted by common infection prevention and control measures, such as hand hygiene.

This report presents a methodology and preliminary results using genetically marked variants of the bacteriophage Lambda (λ) as a harmless surrogate for pathogen transmission. We then validate this method in a naturalistic setting by contaminating different surfaces prior to simulated patient care in a hospital environment.

## Materials and Methods

### Reagents and Equipment

The following reagents and equipment were used in this study: Luria-Bertani Broth Miller (Difco, USA, Product # 244620), LB Plates (Difco, USA, Product # 244510), 0.65% Agar Luria-Bertaini soft agar, LaboPlast Spray Bottle with Pump Vaporizor (Bürkle, Germany, Product # 10216-888), Self-contained 0.85% Saline Swab (Hardy Diagnostic, USA, Product # SRK35), RNase Away Reagent (Invitrogen, USA, Product # 10328-011), DNA AWAY (Molecular BioProducts, Mexico, Product # 7010), 70% ethanol solution (Decon Labs, USA, Product # 2716), Ruler, Disinfecting Wipes (Lysol, China, Product # 3168342), 138 oz. Whirl-Pak (Nasco, USA, Product # B01542), Sterile Saline Wipes (Hygea, USA, Product # C22370), Phusion Blood Direct PCR Master Mix (ThermoFisher, Lithuania, Product # F-175L), O’Gene Ruler DNA Ladder (Thermo Scientific, Lithuania, Product #SM1563), 10,000X GelRed Nucleic Acid Strain (Biotinium, USA, Product # 41003).

### Strains

*Escherichia coli* strain C was acquired from Marie-Agnès Petit from INRAe, France. Bacteriophage λ (λ^Temp^), Bacteriophage λ^Chl^, Bacteriophage λ^Kan^, and Bacteriophage λ^Vir^ was obtained from Maroš Pleška at The Rockefeller University.

### Lysate Preparation

1e^5^ PFU/ml of each phage were cultured with 1e^7^ CFU/ml log-phase *E. coli* C in 10ml of LB broth. These cultures were grown with shaking for six hours before being centrifuged and filtered through a 0.22 μm filter to generate sterile, high-titer lysates of each of the four bacteriophages. These lysates were serially diluted and plated on lawns of *E. coli* C to determine viral titers.

### Phage Distribution

Ten milliliters of each type of λ phage lysate was loaded into their respective spray bottles. The spray bottles were stored at 4°C and transported on ice. Each bottle was primed by spraying five times into a waste container. Immediately after priming, each lysate was sprayed with one pump onto the target site from a distance of 10 cm. Contamination occurred no earlier than thirty minutes before the start of the simulation.

### Phage Recovery

Immediately after the simulation, HCW hands were sampled by applying a saline hand wipe around both hands and forearms. The saline hand wipe was placed into a conical tube for storage. HCWs then placed their disposable scrubs into a Whirl-Pak for storage. High-touch surfaces (See Table I and Figure 1 (11) were sampled with self-contained 0.85% saline swabs; each swab was removed from the saline, and the surface was swabbed in a progressive back and forth motion until the entire surface became damp from the saline. The swab was then returned to the saline solution.

**Table I.** Surfaces sampled in each simulation.

| Type of Surface | Sampling Surface | Number in Figure 1 |
| --- | --- | --- |
| High Touch | Bedside Table | 1,2 (Patient 1); 9,10 (Patient 2) |
|  | Bedrails | 3 (Patient 1); 11 (Patient 2) |
|  | Bedrail Buttons | 4 (Patient 1); 12 (Patient 2) |
|  | Vital Signs Monitor | 7 (Patient 1); 15 (Patient 2) |
|  | Supply Cart | 8 (Patient 1); 16 (Patient 2) |
| Healthcare Worker | Scrubs | 19 |
|  | Bare Hands | 20 |
| WoW | Keyboard | 17 |
|  | Table | 18 |
| Patient Critical Site | Wound | 5 |
|  | Groin | 13 |
| Other Patient Site | Site of Stool Contamination | 6 (Patient 1); 14 (Patient 2) |

**Figure 1.**
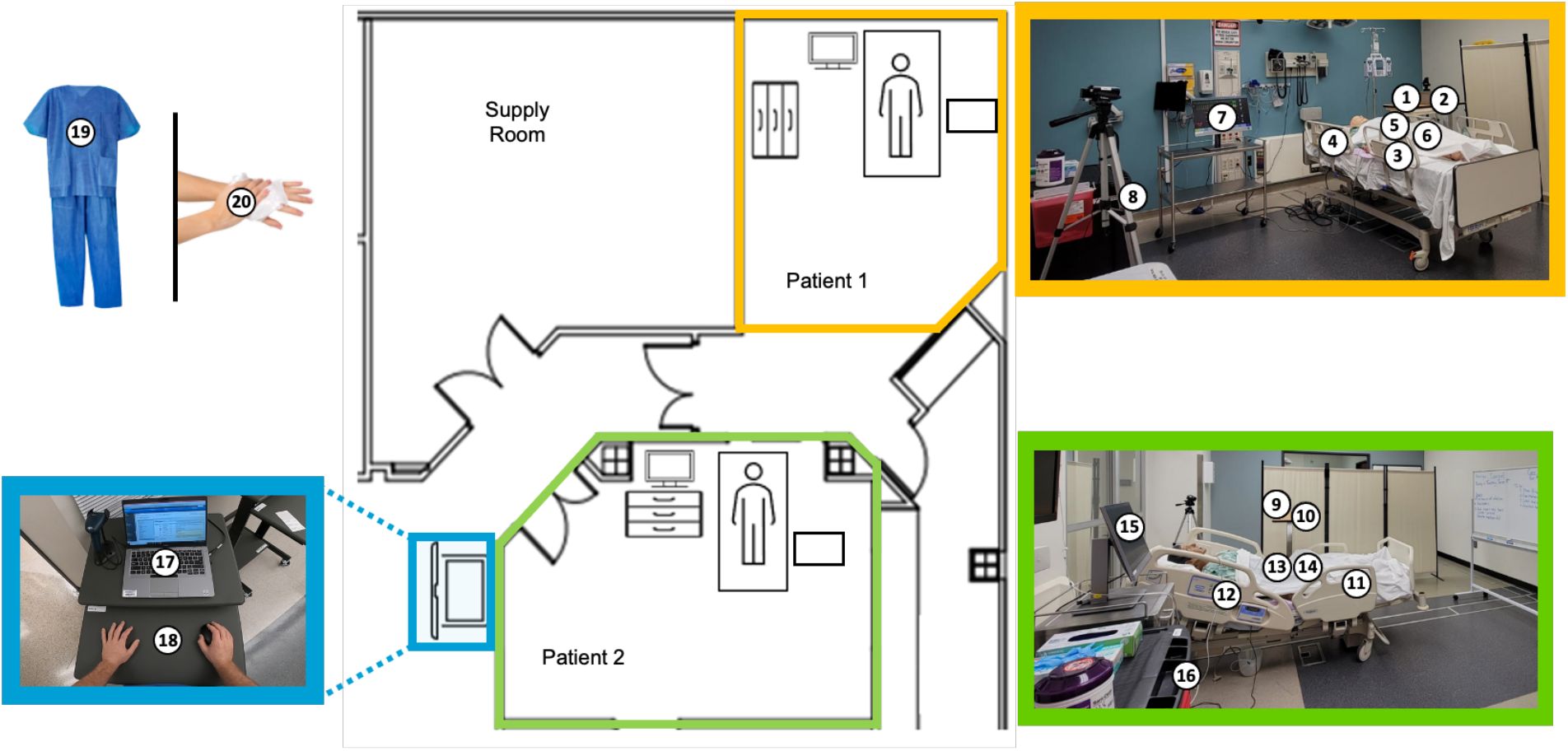
Sampling design. A diagram of the simulated hospital environment with the sampling sites numbered. **(Yellow Inset)** A picture of one simulated hospital room with sampling sites marked. **(Green Inset)** A picture of a second simulated hospital room with sampling sites marked. **(Blue Inset)** A picture of two sampling sites located on the health care worker’s mobile workstation.

### Site Decontamination

After the phage recovery phase, a liberal coating of 70% ethanol was applied to all surfaces upon which the health care worker could have interacted. This alcohol was removed, and this cleaning step was repeated with DNA Away, RNA, and Lysol.

### Phage Identification and Quantification

The samples were tested for the presence, identity, and densities of the four Lambda phages. Phage identification was performed by PCR, using Thermo Scientific’s Phusion Blood Direct PCR Master Mix. Products were visualized on a 1% agarose/TAE gel with Biotium’s 10,000X GelRed Nucleic Acid Stain. Band sizes of 800bp were called λ^Temp^ or λ^Vir^, 1500bp called λ^Chl^, and 1900bp called λ^Kan^. Temperate phage (λ^Temp^) and the virulent mutant (λ^Vir^) were distinguished during the phage quantification step (Supplemental Figure A1).

The serum resistance lipoprotein (*bor*) gene (Gene ID: 2703532, NCBI) of the lambda phages was amplified by PCR using the following primers designed in PrimerBLAST (NCBI): Forward (borRG1Fw) 5’-GCTCTGCGTGATGATGTTGC-3’ and Reverse (borRG1Rv) 5’-GCAGAGAAGTTCCCCGTCAG-3’. Using the double layer soft agar method, LB soft agar overlays containing 0.1 ml of a fully turbid *E. coli C* overnight were prepared and allowed to harden. 0.01 ml of serially diluted saline recovery solution was spotted on the overlay at four densities. These plates were grown overnight at 37°C, and plaques were enumerated the next day. Based on the turbidity of the plaques, λ^Vir^ was distinguished from the other three temperate forms (see Supplemental Figure A2).

### High-Fidelity Simulations

Across two pre-sanitized simulated hospital rooms, defined densities of genetically marked Lambda phages were inoculated at four plausible sources of pathogens on high-fidelity mannequins (e.g., simulated patient excrement, non-intact skin). After contamination, HCWs, comprising registered nurses (RNs) from the emergency department (ED), intensive care units (ICU), or medical/surgical (MS) floors, performed four tasks for two patients over the next hour. For each patient, two of the four tasks required the HCW to interact with a source of contamination: changing a dressing on simulated a stage-4 pressure ulcer (λ^Temp^ on Patient 1’s wound), toileting a patient with a bedpan (λ^Kan^ on Patient 1’s stool), inserting a Foley catheter (λ^Chl^ on Patient 2’s groin), and collecting a stool specimen (λ^Vir^ on Patient 2’s stool). HCWs knew that contamination may be present in the simulation but was unaware of the location of contamination and sampling sites. HCWs wore disposable scrubs over their clothing. Personal protective equipment (e.g., gloves and gowns) and medical-grade disinfectants (e.g., alcohol-based hand rub and disinfectant wipes) were available to every HCW during the simulation. Additionally, HCWs documented their work in an electronic medical record, accessed on a mobile “Workstation on Wheels” (WoW).

## Results

### Method Calibrations

1. To assess the effect of common hospital disinfectants (as well as disinfectants used in simulations; see High-Fidelity Simulations) on λ phages, we tested the ability of different cleaning agents to eliminate the phages independently and in combination and report changes in plaque forming units (Figure 2A). Each disinfectant was able to reduce the amount of bacteriophage by a minimum of three logs and a maximum of four logs. The combination of Ethanol 70%, DNA Away, RNA Away, and Lysol wipes used subsequently on the surface eliminated the phage so that it was undetectable by PCR. The lack of detectability by PCR shows that unlike other biological contamination proxies, such as cauliflower mosaic virus, our method responds to common hospital disinfectants. There was no difference in the reduction ratio to the disinfectants amongst the phages used.

**Figure 2.**
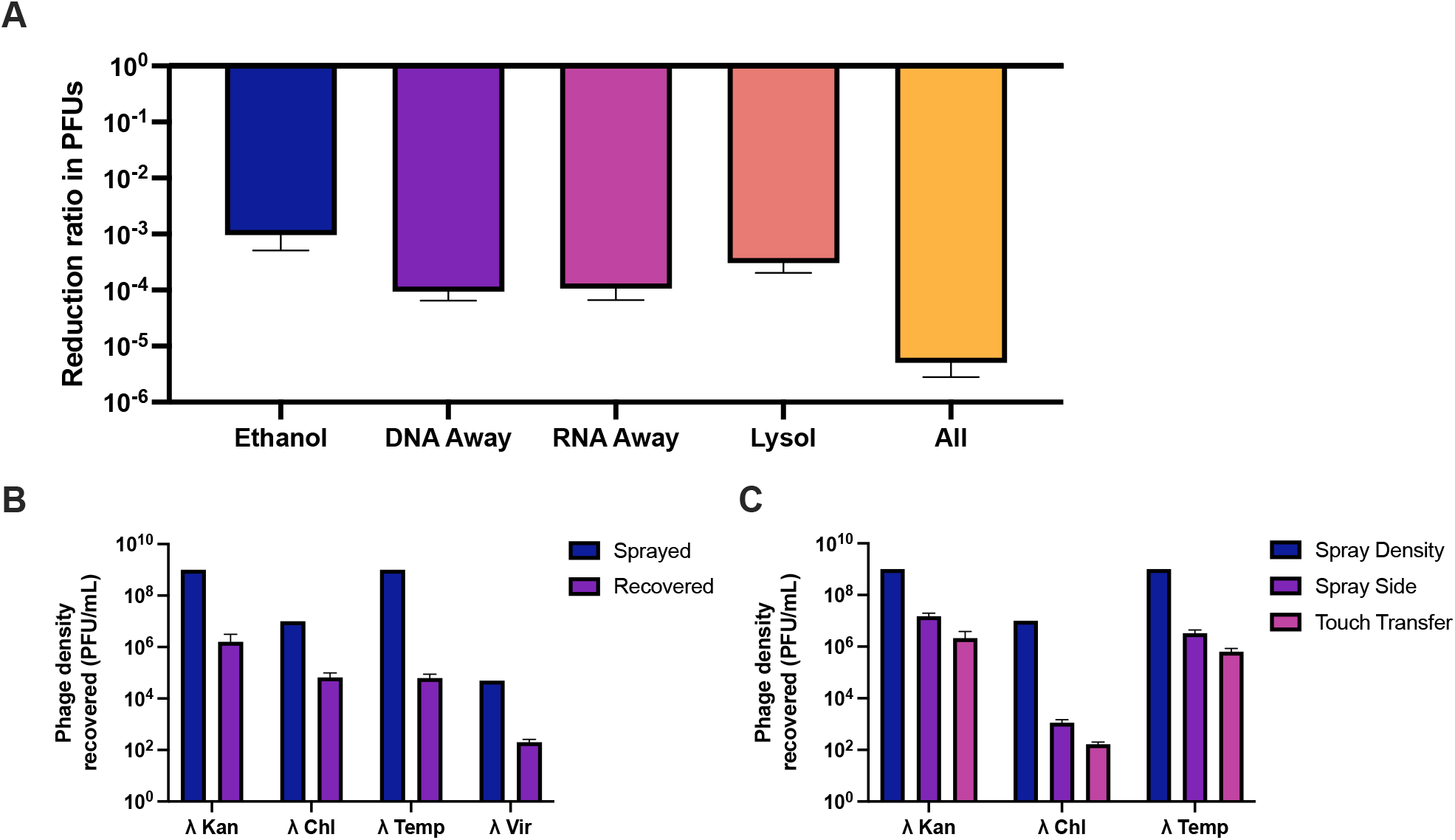
Phage recovery experiments. Experimental results of the effect of antiseptics, time, and surface transfer in sprayed bacteriophage recovery. (**A**) Reduction rate of the phage density on surfaces after exposure to sprayed antiseptics. (**B**) Phage density of the lysate (blue) and the recovery 2 hours post-spray on a surface (purple) at room temperature. (**C**) Phage density of the lysate (blue), the density of the residual phage left on the sprayed surface (purple) after being touched and transferred to a new surface, and the density of phage recovered from the new surface (pink).
2. We estimated the rate of decay of each λ phage variant on a surface over time and from the changes in the density of plaque forming units over a given period (Figure 2B). The bacteriophages were sprayed on a surface and allowed to sit for two hours. The time of two hours was more than the allotted time that the bacteriophages would be sitting on a surface in the medical simulations. After the two hours, the surface was then swabbed and plated for recovery. There was a 2 to 3 log_10_ rate of decay in recoverable PFUs over time. This same rate of decay between lambda phages differentiates it from other bacteriophages used in medical simulations, such as Phi6 and MS2 (6).
3. To determine the efficacy of sampling, phages were sprayed at a known density on a defined area which was then sampled via swabbing. The recovered swabs were then plated on a lawn of *E. coli* C for PFU estimation. We evaluated the efficiency of swabbing recovery to be in excess of 80%.
4. We evaluated the variability between sprays in terms of volume and surface area from a given distance away. To ensure that the spray bottle was distributing a consistent and measurable volume, a weigh boat was placed in an analytical scale and tared. The weigh boat was sprayed with one full squeeze from a spray bottle, and the weight of the sample was recorded. It was assumed that the weight in grams was equivalent to the volume of phage sprayed. The average volume sprayed per pump was 0.215mL (±0.00269). The phage suspension was sprayed from different distances of 5, 10, and 15cm above the bench, to modify the surface area covered. The spray was performed perpendicular to the bench, and the recorded diameter of the spray was defined as the outermost dark ring measured. The diameters for 5, 10, and 15cm away were 65(±1), 80 (±5), and 105 (±10) mm, respectively.
5. To determine the ability of our technique to track phages across multiple surfaces touched by gloved hands, we sprayed a surface with the marked λ phages, which were then touched, spread to other places via subsequent touches, and then sampled. Reported are the changes in plaque forming units between touched surfaces (Figure 2C). After subsequent touching, phage was recovered from the initial inoculation site as well as the site where the phage was transferred via touch. The initial inoculation side saw a one to three-log reduction in PFUs in comparison to the lysate used. The side that was subsequently touched, which initially had no phage, saw a two to four-log reduction in PFUs in comparison to the lysate used. When comparing the initial inoculation side to the subsequently touched side, there was less than a one-log difference in PFUs that were recovered across all lambda phage strains.
6. We evaluated the sensitivity of our PCR detection method bu serially diluting a stock of the marked λ phages and performing PCRs at each dilution (Table I). All strains were 100% recoverable through the 1e^2^ PFU/mL, 58% recoverable (7/12) at 1e^1^ PFU/mL, 58% recoverable (7/12) at 1e^0^ PFU/mL, and 75% recoverable (9/12) at 1e^-1^ estimated PFU/mL. The difference in recovery rate is likely due to chance because of the small sample volume (1uL) used in the PCR processing.

### Method Validation with High Fidelity Simulations

As the high-fidelity simulations were conducted as part of a more extensive study, we present the results of a subset of ten randomly selected simulations performed in the manner described above. For each simulation and λ variant, we measured six binary outcomes: 1) whether transmission of a phage occurred within a patient room, 2) between patient rooms, 3) to the nurse, 4) to the WoW, 5) to a critical site on a patient or 6) to another (non-critical) patient site (Table III). We observed a total of 42 transmission events across the ten simulations, with a median of 5 transmission events (Range = 1 to 8 events) occurring per simulation. Transmission events occurred most frequently within patient rooms (29% of all events), to the nurse (19%), at similar frequencies between patient rooms (14%), to the WoW (14%), or to a critical site on a patient (14%), and least frequently to another (non-critical) site on a patient (10%).

**Table II.**
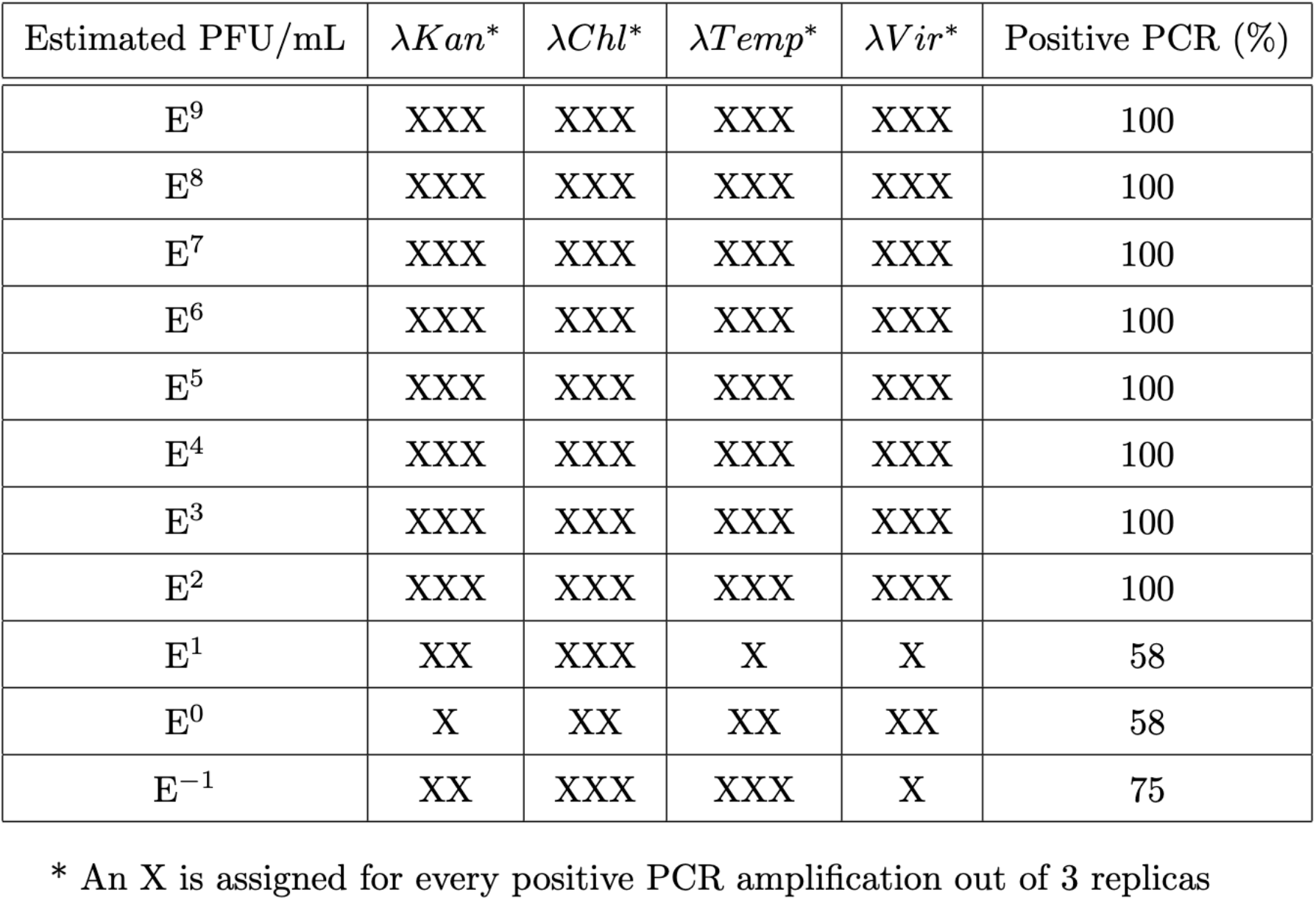
PCR Sensitive for the Detection of Lambda Phage.

| Estimated PFU/mL | $\lambda Kan^*$ | $\lambda Chl^*$ | $\lambda Temp^*$ | $\lambda Vir^*$ | Positive PCR (%) |
| --- | --- | --- | --- | --- | --- |
| E<sup>9</sup> | XXX | XXX | XXX | XXX | 100 |
| E<sup>8</sup> | XXX | XXX | XXX | XXX | 100 |
| E<sup>7</sup> | XXX | XXX | XXX | XXX | 100 |
| E<sup>6</sup> | XXX | XXX | XXX | XXX | 100 |
| E<sup>5</sup> | XXX | XXX | XXX | XXX | 100 |
| E<sup>4</sup> | XXX | XXX | XXX | XXX | 100 |
| E<sup>3</sup> | XXX | XXX | XXX | XXX | 100 |
| E<sup>2</sup> | XXX | XXX | XXX | XXX | 100 |
| E<sup>1</sup> | XX | XXX | X | X | 58 |
| E<sup>0</sup> | X | XX | XX | XX | 58 |
| E<sup>-1</sup> | XX | XXX | XXX | X | 75 |

\* An X is assigned for every positive PCR amplification out of 3 replicas

**Table III.** Definition of transmission events.

| Transmission Event | Definition |
| --- | --- |
| Between-Room | A phage from one patient was recovered from at least one high touch surface in the other patient's room. |
| Within-Room | A phage from one patient was recovered from at least one high touch surface in the same patient's room. |
| Nurse | A phage was recovered from at least one surface on the nurse. |
| WoW | A phage was recovered from at least one surface on the WoW. |
| Patient Critical Site | A phage was recovered from a critical site on a patient, excluding phage introduced to the site as part of the simulation. |
| Other Patient Site | A phage was recovered from a non-critical patient site. |

To illustrate the contribution of each source of contamination to these events, Figure 3 shows the percentage of each type of transmission event originating from each source. All five types of transmission events can be traced back to at least half of the sources of contamination. However, the sources varied in their involvement in transmission events; regarding involvement overall, 50%, 32%, 21%, and 18% of all transmission events originated from Patient 1’s wound (λ^Temp^), Patient 2’s groin (λ^Chl^), Patient 2’s stool (λ^Vir^), or Patient 1’s stool (λ^Kan^), respectively. Regarding involvement in types of transmission events, at least half of transmission events to the nurse (50%) or to the WoW (67%) originated from Patient 1’s wound alone. Most transmission events within patient rooms (83%) originated from either Patient 1’s wound or Patient 2’s stool, and similarly, most transmissions between patient rooms (83%) originated from either Patient 1’s wound or Patient 2’s groin. Patient 1’s stool alone contributed to half of the transmission events to a critical site on a patient. Lastly, phage was infrequently detected on another patient site (i.e., surfaces where the contaminated stool was applied to patients), often originating from another source of contamination on that same patient (e.g., wound or groin). Apart from transmission events, a median of 2 (Range: 1 - 3) of the 4 sources of contamination per simulation was positive for the phage with which they were inoculated, despite each source being cleaned or contained by the HCW.

**Figure 3.**
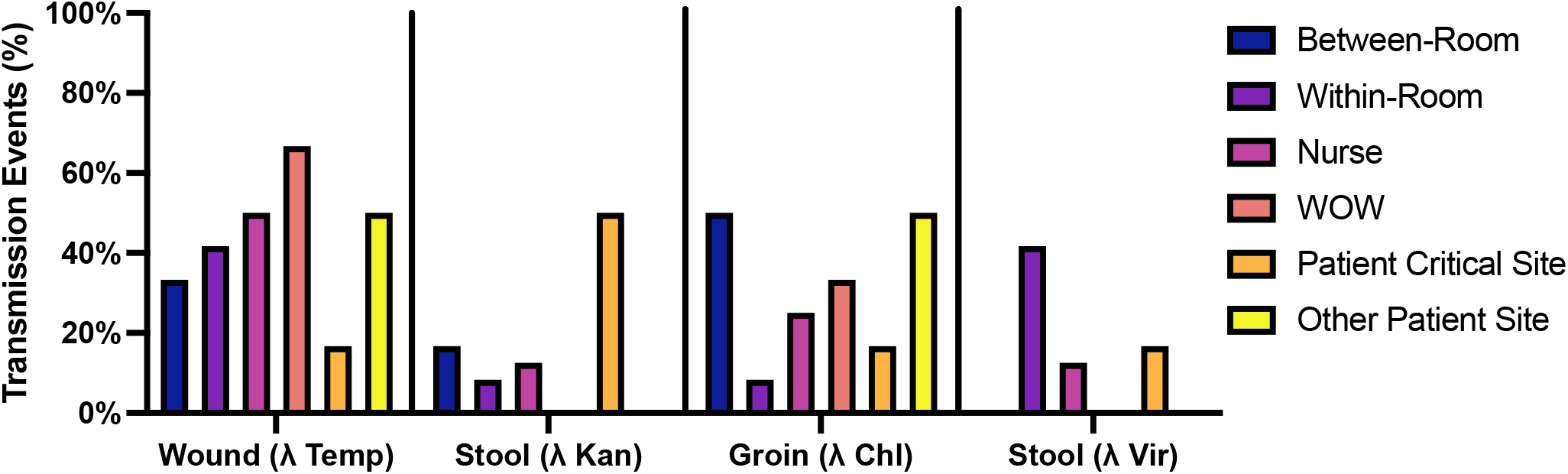
Percentage of transmission event types originating from contamination sources. Shown are data from ten randomly selected simulations. Bars in dark blue are transmissions between the two patient rooms, in orange are transmissions within the patient’s own room, in grey recovery from the nurse, in yellow recovery from the workstation on wheels (WoW), and in light blue recovery of phage from a critical site on a patient. Bars of the same color sum to 100% across the four sources of contamination.

## Discussion

In this report, we describe a method for using naturally occurring variants of bacteriophage λ as surrogates for pathogen transmission. These variants contain unique genetic markers, which permit the identification of the source and transmission path of each phage via PCR. We showed with our calibration experiments that the effect of disinfectants, decay after two hours, and transfer recovery had no difference among the four phages used in this project. The calibrations allow for the interchangeable ability of these viruses if used in different simulated environments and differentiate them from previous methods used to track contaminations.

To validate the transmission dynamics of λ phages in a naturalistic setting, each phage was inoculated onto a surface in a simulated hospital environment prior to simulated patient care. The patterns of dissemination observed in these simulations resemble those that occur during actual patient care; in the simulations, most transmission events involved the movement of phage from a patient to a high touch surface in that patient’s room (e.g., to the bedrails, bedside table, or vital signs monitor). In clinical practice, frequent contact between a HCW, their patient, and the patient’s immediate environment rapidly colonizes high-touch surfaces with a patient’s own flora (12). Moderately frequent were transmission events to the HCW (e.g., clothing or hands), which contribute to transmission between patients or their rooms (10). Among the least frequent events but most concerning was the transmission of phage to a critical site on a patient. These events are relatively uncommon during actual patient care (2) but increase the risk of a patient developing an infection (12).

An advantage of the present method is that it allows for transmission events to be traced back to different sources of contamination (e.g., patient care tasks). Demonstrating that the four phages were differentially involved in transmission events in the simulations provides further validation of their dynamics in naturalistic settings; for example, phage from Patient 1’s wound (a simulated stage-4 pressure ulcer) was the variant most frequently involved in transmission events, particularly transmission to the WoW or the nurse. Wound care is considered a high-contact patient care task, which creates opportunities for pathogens to be transferred to HCW hands or clothing (13). Consequently, in 2019, the Centers for Disease Control and Prevention began recommending the use of gowns and gloves when performing wound care in skilled nursing facilities (where multidrug-resistant organisms (MDRO) transmission is common), regardless of a resident’s MDRO colonization or infection status (13). In accordance with this recommendation, the present method identified wound care as a frequent contributor to transmission events, particularly those that may disseminate pathogens between patients.

As calibration experiments demonstrated the equivalence of the λ variants, differences in how each variant was disseminated in simulated patient care reflect other factors, such as characteristics of tasks (e.g., amount of patient contact) or the infection prevention practices of HCWs. Consequently, this method is useful for identifying the sources of contamination that contribute most to transmission events, where sources could be located simultaneously in different rooms (e.g., to examine MDRO transmission between patients; (10)), body sites of a patient (e.g., to assess the role of endogenous microbes in hospital-acquired infections; (12)), or surfaces on a HCW (e.g., to evaluate the contribution of different items of PPE to HCW self-contamination during doffing;(7)). Lastly, unlike similar surrogates (9), λ phages are susceptible to common disinfectants like alcohol-based hand rub and surface disinfectants, so the effect of infection prevention practices on transmission may also be evaluated.

The present method is not without limitations. Unlike other surrogates, like fluorescent tracers, live viruses do not provide immediate feedback about the occurrence of transmission events. Although less useful for rapidly training HCWs in infection prevention practices (7), the ability to simulate multiple sources of pathogens simultaneously and realistically lends itself to rigorous research or quality improvement efforts (e.g., evaluating the effectiveness of the training program). Lastly, the number of available λ variants is limited, but any similar phages with detectable variation at one gene could be employed for future efforts.

## Conclusions

Four variants of the bacteriophage λ were used as surrogates for pathogens to track transmission events in a simulated hospital environment. Analyses of the results from ten simulations in which healthcare workers performed common patient care tasks revealed that λ phages can identify the sources of pathogen transmission and assess their differential involvement in transmission events within and between patient rooms, to mobile surfaces, and to critical sites on patients. Whereas existing approaches using non-biological or biological surrogates for pathogens have succeeded in simulating multiple sources of contamination but not the susceptibility of pathogens to common disinfectants (e.g., alcohol-based hand rub), the present method is notable for achieving both. The applicability of the present method is broad but is particularly relevant to understanding the sources and pathways of MDRO transmission in healthcare settings.

## Data Availability

The data supporting this study's findings are available in this manuscript and supplementary
material. Further requests may be made to the corresponding author.

## Acknowledgments

We would like to thank Rebecca MacKay for her comments and support in the simulations, Jill Morgan, Golpar Ghassemian, and Lindsey Lee for their help designing the simulation scenario, and Ellen Jordan, Anagha Nair, and Marilyn Garcia for conducting the simulations. We would also like to thank the nurses who participated in the simulations.

BRL would like to thank the US National Institutes of General Medical Sciences for their funding support via R35 GM 136407.

## Data Availability

The data supporting this study’s findings are available in this manuscript and supplementary material. Further requests may be made to the corresponding author.

## Strain Availability

All bacterial strains and bacteriophages used in this project are available upon request to Joel Mumma or Bruce Levin.

