## Supplemental Materials for "Using Lambda phages as a proxy for pathogen transmission in hospitals"

**Supplementary Information**

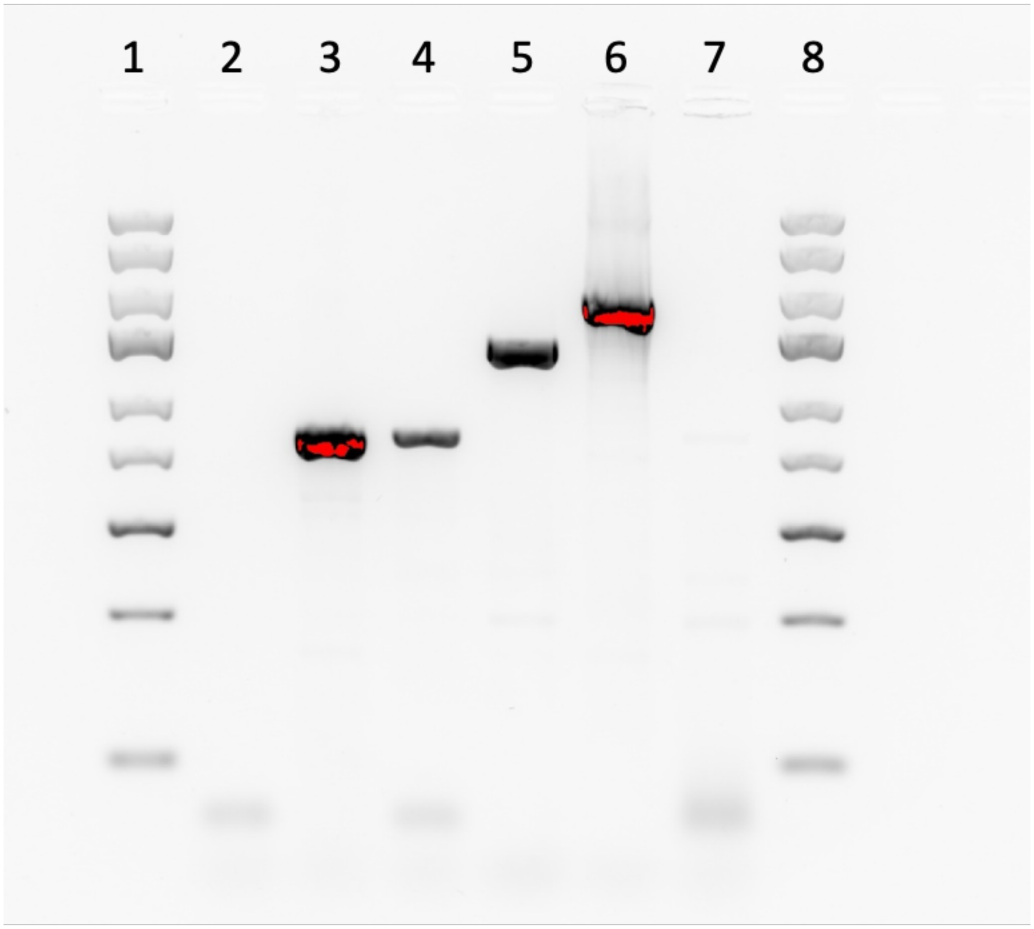


**Supplemental Figure A1. PCR Differentiation of Lambda phage variants.** Shown from left to right are the (1) Standard Ladder, (2) a water control, (3) $\lambda^{Temp}$, (4) $\lambda^{Vir}$, (5) $\lambda^{Chl}$, (6) $\lambda^{Kan}$, an (7) *Escherichia coli* bacterial host control, and a (8) O'GeneRuler Express DNA Ladder (5kb).

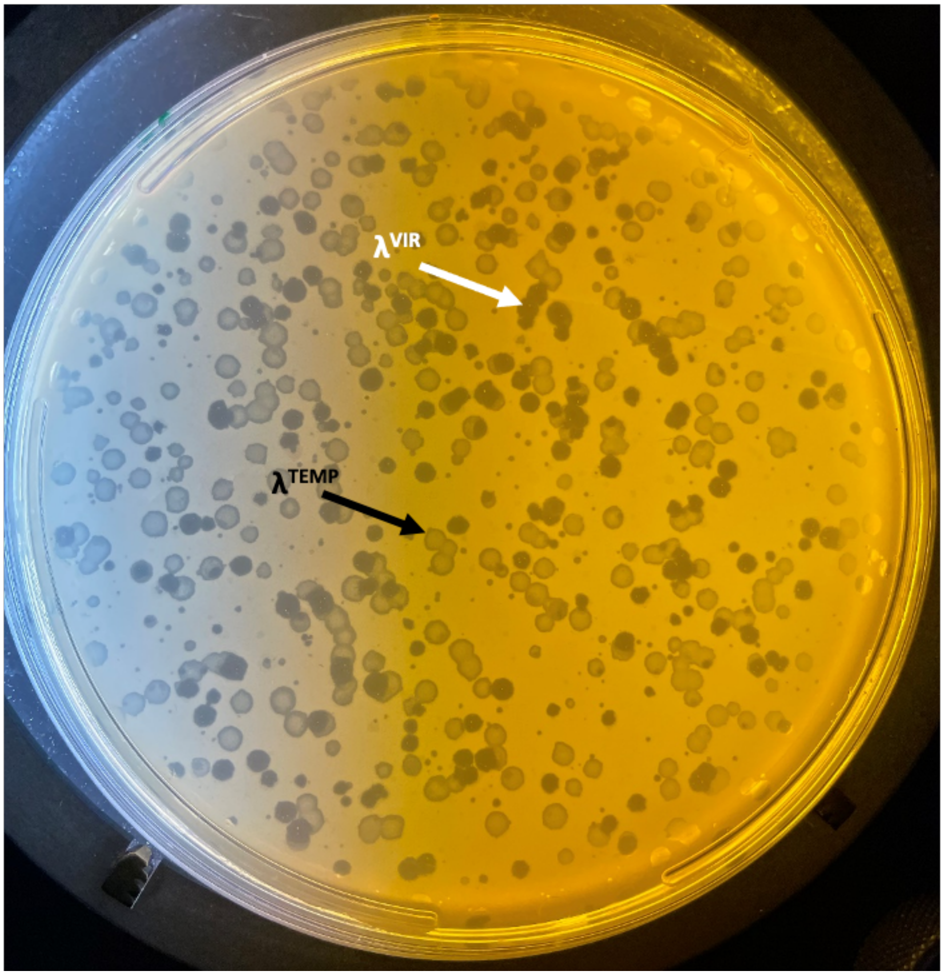


**Supplemental Figure A2. Plaque morphology difference in $\lambda^{Temp}$ and $\lambda^{Vir}$.** Shown is a double-layer soft agar lawn of *E. coli* C containing both $\lambda^{Temp}$ (turbid plaques) and $\lambda^{Vir}$ (clear plaques).
